# Vaccine preparation time, errors, satisfaction, and preference of prefilled syringes versus RSV vaccines requiring reconstitution: randomized, time and motion study

**DOI:** 10.1101/2024.04.16.24305921

**Authors:** Darshan Mehta, Samantha Kimball-Carroll, Dayna R. Clark, Serena Fossati, Matthias Hunger, Ankit Pahwa, Mia Malmenas, Brian Hille, Nicolas Van de Velde

## Abstract

**Introduction:** RSV infections can lead to serious outcomes, especially among older adults. Two United States (US) Food and Drug Administration (FDA) approved vaccines, both requiring reconstitution (VRR) prior to administration, are recommended by the Advisory Committee on Immunization Practices (ACIP) for adults aged 60+ years. An alternative vaccine employing a ready-to-use prefilled syringe (PFS) is currently under evaluation by the FDA. The current study compared a PFS versus two VRRs (VRR1 and VRR2) to evaluate preparation time, errors, satisfaction, and preference in a randomized, single-blinded time and motion (T&M) study.

**Methods:** Participants were recruited and randomized to a preparation sequence of the three vaccines. Participants read instructions, then consecutively prepared the three vaccines with a 3- to 5-minute washout period in between. Preparations were video recorded and reviewed by a trained pharmacist for preparation time and errors using predefined, vaccine-specific checklists. Participant demographics, satisfaction with vaccine preparation, and vaccine preference were recorded. Within-subjects analysis of variance (ANOVA) was used to compare preparation time. Mixed-effects Poisson and ordered logistic regression models were used to compare number of preparation errors and satisfaction scores, respectively.

**Results:** 63 pharmacists (60%), nurses (35%), and pharmacy technicians (5%) participated at four sites in the US. The least squares (LS) mean preparation time per dose for PFS was 141.8 seconds (95% CI:156.8, 126.7; p<0.0001) faster than for VRR1, 103.6 seconds (118.7, 88.5; p<0.0001) faster than for VRR2, and 122.7 seconds (95% CI: 134.2, 111.2; p<0.0001) faster than the pooled VRRs. Overall satisfaction (combined ‘Very’ and ‘Extremely’) was 87.3% for PFS, 28.6% for VRR1, and 47.6% for VRR2. Most participants (81.0%) preferred the PFS vaccine.

**Conclusion:** PFS vaccines can greatly simplify the vaccine preparation process, allowing administrators to prepare almost four times more doses per hour than with vial and syringe systems.

**Key Summary Points:** *Why carry out this study?:*

- Two US FDA approved vaccines against RSV require reconstitution. An alternative vaccine employing a ready-to-use prefilled syringe (PFS) is currently under evaluation by the FDA. **●** We conducted the current study to compare the impact of RSV vaccine format on preparation time, errors, satisfaction and preference between a ready-to-use single-dose prefilled syringe (PFS) RSV vaccine versus two RSV vaccines requiring reconstitution (VRRs).

*What was learned from the study?:*

- Preparation time with PFS was reduced by a factor of 4 compared to VRRs.
- Most healthcare professionals were extremely satisfied and preferred a PFS presentation over VRRs.
- PFS vaccines can help vaccine administrators save time on preparation resulting in nearly quadruple their hourly vaccine preparation rate compared to VRRs.

## Introduction

Respiratory syncytial virus (RSV) is a common respiratory virus, causing approximately 159,000 hospitalizations, 119,000 emergency department admissions, and 1.4 million outpatient visits annually in US adults aged 65 years and older [1]. Although adjusted for under-detection due to imperfect sensitivity of reverse transcription polymerase chain reaction testing of nasopharyngeal or nasal swabs, these figures are likely still underestimated for several reasons, including, but not limited to, the lack of routine RSV testing in clinical practice [1].

RSV usually causes mild, cold-like symptoms, but it can cause serious illness in adults (≥65 years) and infants [2]. Compared to coronavirus disease (COVID-19) or influenza, older adults hospitalized with RSV often have more severe disease, are more likely to be admitted to the intensive care unit, and receive standard flow oxygen, high-flow nasal cannula, or noninvasive ventilation [3]. After hospital discharge, up to 15% of these individuals require a higher level of care than needed prior to admission [4, 5]. Infection with RSV is costly, with an estimated annual economic burden in US adults (≥60 years) of $6.6 billion, with $2.9 billion in direct medical costs, $1.1 billion in indirect costs due to losses in productivity from RSV-related morbidity, and $2.5 billion in indirect costs due to RSV mortality [6].

Advisory Committee on Immunization Practices (ACIP) currently recommends that adults over 60 years may receive a single dose of RSV vaccine, using shared clinical decision-making with their healthcare provider (HCP) [7]. Two RSV vaccines are currently approved by the US Food and Drug Administration (FDA) for adults aged 60 years and older – AREXVY® (RSVPreF3, GSK) and ABRYSVO™ (RSVpreF, Pfizer), both of which are vaccines requiring reconstitution (VRR). AREXVY® (from here on addressed as VRR1) includes two vials, one with a lyophilized vaccine (powder) and one with an adjuvant suspension. Preparation steps for VRR1 include cleansing the vial stoppers, transferring the diluent into the vial of lyophilized vaccine using a separate syringe and needle, swirling to dissolve the powder, and withdrawal of the reconstituted vaccine into the syringe for administration [8]. ABRYSVO™ (addressed as VRR2) is available in two presentations: i) two vials with one vial of lyophilized vaccine and one vial of liquid diluent, and ii) one vial with lyophilized vaccine and a pre-filled syringe (PFS) with sterile water diluent. Preparation of VRR2 in the second format, as utilized in the current study, includes cleansing the vial stopper, attachment of the provided adapter to the vial, connection of the PFS to the vial adapter, transferring the diluent from the PFS into the vial of lyophilized vaccine, swirling to dissolve the powder, withdrawal of the reconstituted vaccine for administration into the PFS, and locking the needle [9].

Moderna is currently developing an mRNA-based RSV vaccine (mRNA-1345), under review with the FDA, which employs the same ready-to-use prefilled syringe (PFS) delivery system used to administer the FDA-approved SPIKEVAX® (mRNA-1273, Moderna) (addressed as PFS). Preparation of PFS includes removing the PFS tip cap and locking the needle (needle not included in the vaccine kit) [10]. Preparation of mRNA-1273 PFS was utilized in the current study as a proxy for mRNA-1345.

Different vaccine presentations involve varying time and risks for errors during preparation, both factors contributing to overall cost. In comparison with VRR, PFS are generally easier to handle and administer, and are associated with reductions in both time and errors in preparation [11, 12]. De Coster et al. conducted a cross-over, randomized, open-label time and motion (T&M) study, and recorded HCPs as they prepared one PFS and one VRR vaccine consecutively with a 3-to-5-minute washout period in between the preparations [13]. Overall, PFS preparation resulted in a mean time saving of 34.5 sec [95% CI 28.4; 40.6] compared to VRR, and out of 96 preparations of each presentation, there were 10 (10.4%) errors reported during PFS preparation compared to 47 (48.9%) for VRRs [13]. Another study indicated an average time saving of 1.1 min (66 sec) for each PFS prepared and administered vaccine dose compared to VRR [11].

In the selection of vaccine presentations, HCP preference may play a role. Several studies have reported higher HCP preference for PFS presentations, with the most common reasons being reduced number of immunization errors, ease of administration, and work efficiency [11, 14–17]. HCPs also expressed that using PFS lowers the risk of needle contamination and needle stick injuries and reduces the possibility of dosage errors [15, 17].

Up to now, studies have not addressed preparation time and errors along with satisfaction and preference of HCPs, nor have they been conducted with vaccines specific to RSV. T&M studies include independent and continuous observation and are more precise than self-reporting or work sampling techniques, which collect data at intervals of time. This study design has been used in healthcare to determine the timing and duration of procedures or tasks, and to calculate vaccine preparation time and errors [13, 18]. Video recording of HCPs has been shown to reduce the observer effect and has the potential to improve the quality of data collected by providing the opportunity to replay each task for review and analysis.

### Study Objectives

The primary objective of the study was to assess and compare the vaccine preparation times of PFS, VRR1, and VRR2 prior to administration. Secondary objectives of the study were to assess and compare the rate of vaccine preparation errors, participant satisfaction, and preference for PFS, VRR1, and VRR2.

We hypothesized that compared to VRR1 and VRR2, PFS would result in reduced preparation time, fewer errors, and higher HCP satisfaction and preference.

## Methods

### Study design

A randomized, 3-period, 3-sequence cross-over, single-blinded T&M study was conducted at four sites in the United States (US) (**Figure 1**). To maintain single-blind for the study, the PFS presentation was labeled ‘Vaccine A’, VRR1 was labeled ‘Vaccine B’ and VRR2 was labeled ‘Vaccine C’. The recruited participants were randomly assigned a vaccine preparation sequence (ABC, BCA, or CAB) blocked by site for balance. On arrival at the site, participants were asked to read blinded vaccine preparation instructions for each of the three study vaccines (generated from package inserts for each vaccine [8–10]), and to complete an electronic survey on site (**Supplementary material 1**) to gather their demographic information.

**Figure 1.**
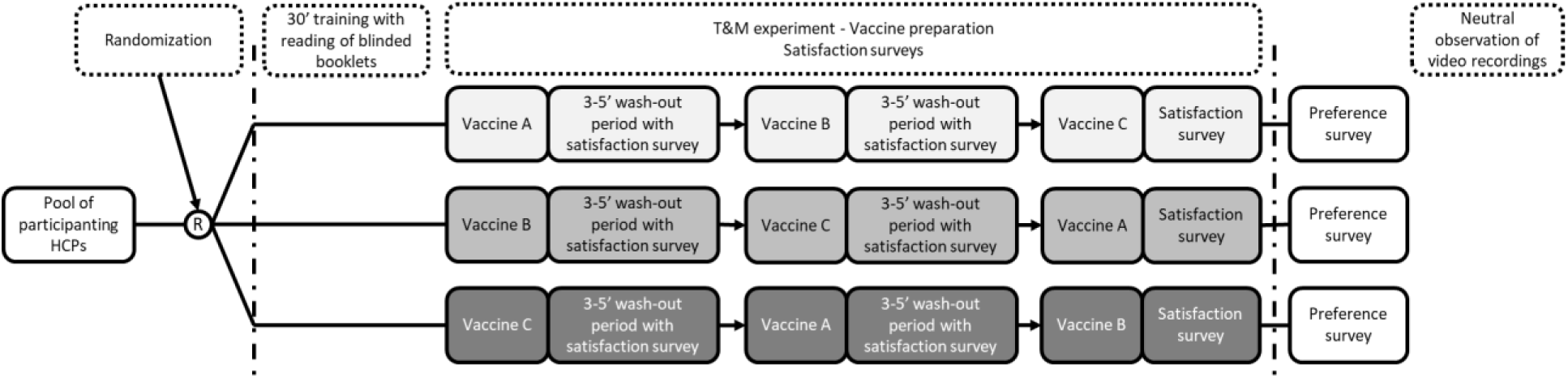
Study design. Abbreviations: HCPs, healthcare providers; Vaccine A, PFS; Vaccine B, VRR1; Vaccine C, VRR2.

After a period of approximately 30 minutes, participants were asked to consecutively prepare one dose of each vaccine in the assigned randomized sequence with a 3- to 5- minute washout period in between each preparation. Vaccines labels were blinded, and vaccines were laid out on the counter for participants, along with required materials to facilitate aseptic technique as per usual practice. During each washout period, the participants rated their satisfaction with the vaccine ease of preparation, preparation time and overall satisfaction. After preparing all 3 vaccines, participants provided their overall preference between vaccine A, B or C.

Vaccine preparations were video recorded on-site by trained nurses. An independent, experienced, trained pharmacist reviewed each video to assess the time taken for each preparation, and documented any preparation errors using predefined, vaccine-specific checklists. A second pharmacist independently reviewed 24% (first nine participants and six randomly selected participants) preparation videos as quality control. Agreement between reviewers of ≥80% was considered acceptable, and no changes were needed based on quality control. The vaccine-specific error checklist was developed by reviewing all steps listed in the label of approved vaccines and were cross referenced with published literature [13, 16].

On-site study personnel were trained to avoid any influence on the participant during vaccine preparation. Vaccine preparation was conducted in a research setting mimicking that of a busy retail pharmacy/clinic (i.e., open area with background noise from clinic daily activities). Study processes, including the completeness of error checklists, underwent internal validation with the first three study participants.

### Sample size

De Coster et al. reported average preparation time for VRR of 70.5 seconds and for PFS of 36.0 seconds, with a mean difference [SD] of 34.5 [30] seconds, i.e., roughly half the time [13]. This suggests a large effect size (>0.5) [19]. For the sample size calculations, we used a 0.5 effect size and a paired t-test, resulting in a samples size of 54 (18 for each cohort) with a statistical power of 90% at α=0.025. Applying a non-parametric correction of 15% and assuming a 10% drop-out rate, the sample size increased to 63 (21 for each of the three sequence cohorts) [20].

### Participants

Participants were eligible if they were at least 21 years of age, had a minimum 1.5 years of experience administering vaccines, had administered at least one vaccine in the month prior, and were either one of the following: US-licensed and practicing pharmacists, US-registered practicing nurse, or a practicing pharmacy technician certified by the Pharmacy Technician Certification Board. Participants with any current business relationship with Moderna, Pfizer, and/or GSK were excluded, including, but not limited to, employment, consultancy agreement, and/or holding individually managed stocks. The participants were screened into the study based on self-report of the inclusion and exclusion criteria.

### Participant recruitment

Recruiters were trained to conduct recruitment in accordance with study criteria. The recruitment process involved outreach to an expert network of pharmacists, pharmacist technicians, and nurses.

Further potential participants were also identified via email, LinkedIn, cold calls, and referrals. The largest source of participants came via LinkedIn and the second largest through referrals.

### Outcomes

The outcome for the primary objectives was vaccine preparation time, defined as the time in seconds from when the participant touched any material on the experimental field (e.g., syringe, vial, needles) to the time the syringe ready for administration was laid down on the table.

Secondary outcomes included the total number of vaccine preparation errors and participants’ vaccine satisfaction and preference. The total number of errors was defined as the sum of specific errors for each participant and vaccine recorded by the observer in the vaccine-specific checklists. Observer comments made in the ‘other’ error category were reviewed by the study team. Comments were not considered ‘other’ errors if they were not mistakes made by the participant during preparation (i.e. preparation was out of view of the camera for a moment), were errors that were already accounted for by another error indicated by the observer in the checklist, or if they could reasonably be incorporated into an existing error category. Further, repetitive errors identified in the others category were pulled out into a new category (e.g., used more than one syringe). See **Supplementary materials 2** and **3** for more details on errors.

Satisfaction for ease-of-preparation, time taken for preparation, and overall vaccine preparation procedure were assessed through a self-administered online survey using a 7-point Likert scale. Satisfaction for each outcome was measured on a 7-point Likert scale (Extremely Dissatisfied; Very Dissatisfied; Dissatisfied; Neither Satisfied nor Dissatisfied; Satisfied; Very Satisfied; Extremely Satisfied). Vaccine preference was determined through a single item where participants selected the vaccine they preferred most.

### Ethics, data privacy and pharmacovigilance

This study was conducted in compliance with the principles of the Declaration of Helsinki. The protocol and each study site were reviewed and approved by a central institutional review board (Advarra).

Written informed consent was obtained from each participant prior to starting the study. In this study no vaccine was administered. Participants who completed the study received fair market value (FMV) compensation for their time. Their travel time was compensated for half the hourly FMV rate for a maximum of 1.5 hours.

### Statistical analysis

To estimate mean preparation time (seconds) for each vaccine and to test the difference in vaccine preparation times, a within-subjects analysis of variance (ANOVA) approach was implemented through a linear mixed model (LMM). For the pairwise comparison of individual vaccines, the Dunnett multiple comparison adjustment was used. Normality of residuals was confirmed using quantile-quantile (q-q) plots. Comparisons of the number of preparation errors made for each vaccine was performed using a Poisson generalized linear mixed model (GLMM). The potential presence of over- (or under-) dispersion by fitting a negative binomial GLMM and assessing the significance of the dispersion parameter k. Comparisons of the satisfaction scores between vaccines were performed using mixed-effects ordered logistic regression models.

All models included preparation sequence and vaccine (i.e. PFS, VRR1, and VRR2) as fixed effects, and random effects (random intercepts) for subject and site. For endpoints related to preparation time, supplementary models were run including participant career experience (years), experience administering vaccines (years), and occupation as fixed effects.

Post-hoc analyses were run comparing PFS to data from pooled VRRs (pVRR: data from VRR1 and VRR2 combined into a single cohort) for all endpoints. A p-value lower than 0.05 was considered statistically significant. All analyses were conducted using SAS v9.4.

### Participant recruitment and characteristics

Recruitment involved outreach to 1,069 individuals, of which 80 were deemed eligible, and a total of 63 participants were enrolled and completed the study (**Figure 2**).

**Figure 2.**
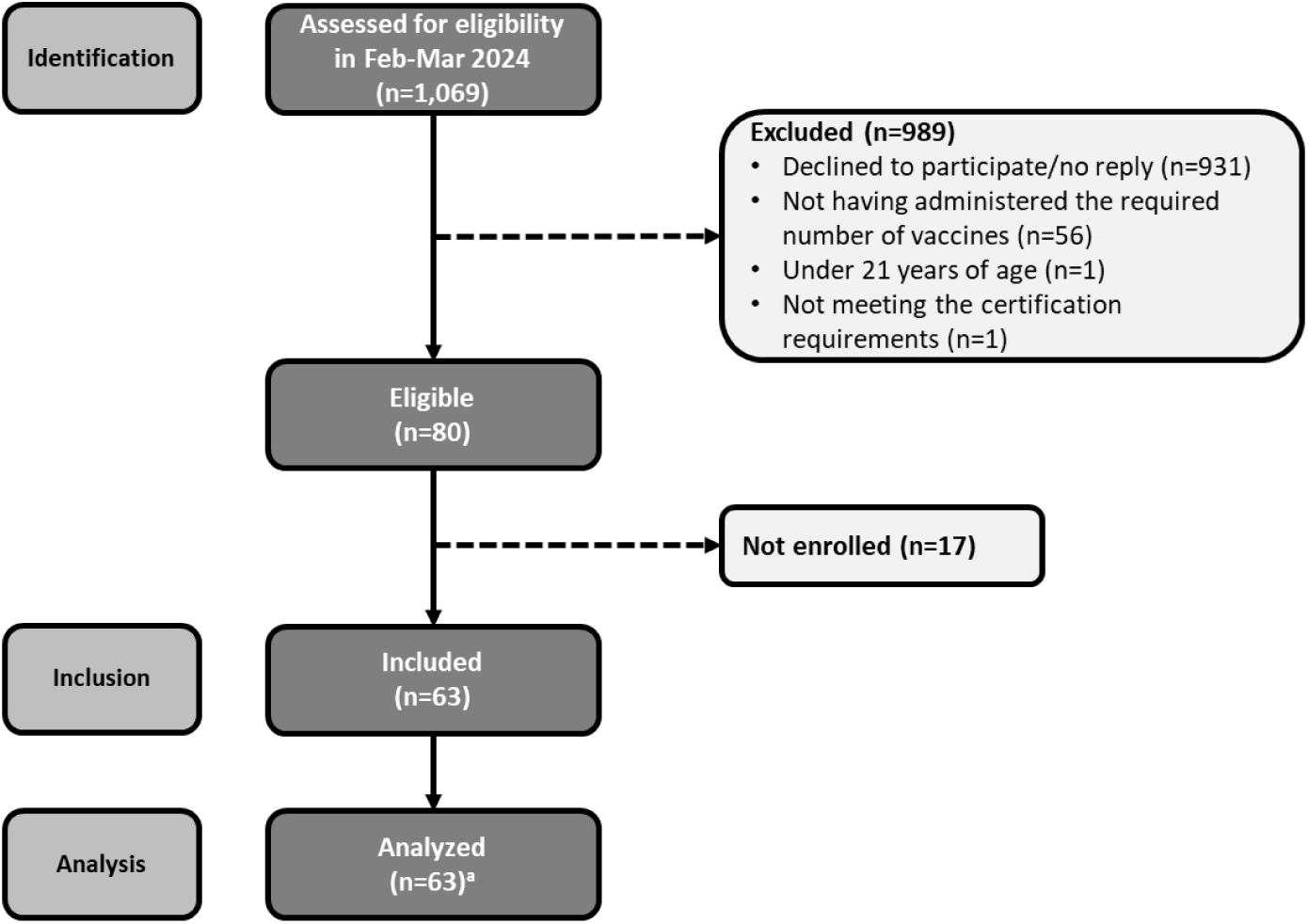
STROBE flow chart.

Demographics of the 63 participants who completed the study (21 randomized to each of the 3 vaccine preparation sequences) are presented in **Table 1**. The mean (SD) age of participants was 41.9 years (10.7) and 31.7% were male (n=20). The majority were either pharmacists (n=38, 60.3%) or nurses (n=22, 34.9%). Participants were largely from urban practices (n=47, 74.6%), and primarily worked in a retail pharmacy setting (n=36, 57.1%). Most participants had less than 20 years of experience in their current occupation, with 39.7% (n=25) having between one and 10 years of experience, and 33.3% (n=21) having between 11 and 20 years of experience. Over half had more than 10 years of experience administering vaccines (n=32, 50.8%). Almost all indicated general training on vaccine preparation (n=61, 96.8%) and specific training for VRR preparations (n=60, 95.2%), while specific training for PFS preparations was less common (n=53, 84.1%).

**Table 1.** Participant demographics, overall and by randomization sequence.

|  | Overall<br>N=63 | Sequence ABC<br>N=21 | Sequence BCA<br>N=21 | Sequence CAB<br>N=21 |
| --- | --- | --- | --- | --- |
| **Age (years), mean (SD)** | 41.9 (10.7) | 41.5 (11.0) | 44.2 (10.8) | 39.9 (10.5) |
| **Male, n (%)** | 20 (31.7) | 7 (33.3) | 6 (28.6) | 7 (33.3) |
| **Occupation, n (%)** |  |  |  |  |
| Pharmacist | 38 (60.3) | 14 (66.7) | 13 (61.9) | 11 (52.4) |
| Nurse | 22 (34.9) | 6 (28.6) | 8 (38.1) | 8 (38.1) |
| Pharmacy technician | 3 (4.8) | 1 (4.8) | 0 (0.0) | 2 (9.5) |
| **Practice, n (%)<sup>1</sup>** |  |  |  |  |
| Urban | 47 (74.6) | 14 (66.7) | 16 (76.2) | 17 (81.0) |
| Suburban | 21 (33.3) | 6 (28.6) | 8 (38.1) | 7 (33.3) |
| Rural | 5 (7.9) | 2 (9.5) | 0 (0.0) | 3 (14.3) |
| **Practice setting, n (%)<sup>1</sup>** |  |  |  |  |
| Retail pharmacy | 36 (57.1) | 14 (66.7) | 12 (57.1) | 10 (47.6) |
| Hospital | 27 (42.9) | 4 (19.0) | 12 (57.1) | 11 (52.4) |
| Academic institution | 8 (12.7) | 3 (14.3) | 2 (9.5) | 3 (14.3) |
| Clinical pharmacy | 5 (7.9) | 0 (0.0) | 2 (9.5) | 3 (14.3) |
| Other | 11 (17.5) | 4 (19.0) | 1 (4.8) | 6 (28.6) |
| **Years experience, n (%)** |  |  |  |  |
| 1-10 years | 25 (39.7) | 9 (42.9) | 6 (28.6) | 10 (47.6) |
| 11-20 years | 21 (33.3) | 7 (33.3) | 8 (38.1) | 6 (28.6) |
| 21-30 years | 9 (14.3) | 2 (9.5) | 4 (19.0) | 3 (14.3) |
| 31-40 years | 4 (6.3) | 1 (4.8) | 2 (9.5) | 1 (4.8) |
| >40 years | 4 (6.3) | 2 (9.5) | 1 (4.8) | 1 (4.8) |
| **Average number of vaccines administered in a typical month, n (%)** |  |  |  |  |
| 1-10 vaccines | 25 (39.7) | 7 (33.3) | 11 (52.4) | 7 (33.3) |
| 11-20 vaccines | 8 (12.7) | 4 (19.0) | 0 (0.0) | 4 (19.0) |
| 21-30 vaccines | 13 (20.6) | 4 (19.0) | 4 (19.0) | 5 (23.8) |
| 31-40 vaccines | 5 (7.9) | 0 (0.0) | 2 (9.5) | 3 (14.3) |
| >40 vaccines | 12 (19.0) | 6 (28.6) | 4 (19.0) | 2 (9.5) |
| **Years administering vaccines, n (%)** |  |  |  |  |
| 1.5-3 years | 8 (12.7) | 2 (9.5) | 2 (9.5) | 4 (19.0) |
| >3-5 years | 10 (15.9) | 4 (19.0) | 2 (9.5) | 4 (19.0) |
| >5-10 years | 13 (20.6) | 4 (19.0) | 4 (19.0) | 5 (23.8) |
| >10 years | 32 (50.8) | 11 (52.4) | 13 (61.9) | 8 (38.1) |
| **Training<sup>1</sup>, n (%)** |  |  |  |  |
| Vaccine preparation | 61 (96.8) | 19 (90.5) | 21 (100.0) | 21 (100.0) |
| VRR preparation | 60 (95.2) | 18 (85.7) | 21 (100.0) | 21 (100.0) |
| PFS preparation | 53 (84.1) | 16 (76.2) | 17 (81.0) | 20 (95.2) |
| **Experience<sup>1</sup>, n (%)** |  |  |  |  |
| Administering VRR | 60 (95.2) | 19 (90.5) | 20 (95.2) | 21 (100.0) |
| Administering PFS | 58 (92.1) | 19 (90.5) | 18 (85.7) | 21 (100.0) |

Abbreviations: A, PFS; B, VRR1; C, VRR2; PFS, prefilled syringe; SD, standard deviation; VRR, vaccine requiring reconstitution. <sup>1</sup> The sum of the percentages of individual categories may exceed 100% because multiple selection was allowed for this question.

### Vaccine preparation time

Complete vaccine preparation time were available for all participants for PFS and VRR1, and for 62 participants for VRR2, due to the malfunctioning of the vaccine kit vial adapter for one participant who was unable to complete the preparation (**Table 2**). Correlation of preparation time between observers was high (Pearson’s r=0.99, n=15). The observed mean time (SD) for vaccine preparation was 43.5 seconds (23.8) for PFS, 185.3 seconds (57.1) for VRR1, 147.2 seconds (56.9) for VRR2, and 166.4 seconds (59.9) for the pooled VRR cohort (pVRR).

**Table 2.** Vaccine preparation time, by vaccine and for pooled VRRs, with comparison.

|  | PFS | VRR1 | PFS vs. VRR1<sup>1,2</sup> | VRR2<sup>3</sup> | PFS vs. VRR2<sup>1,2</sup> | pVRRs<sup>3</sup> | PFS vs. pVRRs<sup>1</sup> |
| --- | --- | --- | --- | --- | --- | --- | --- |
| **Time for vaccine preparation (seconds)** |  |  |  |  |  |  |  |
| N | 63 | 63 |  | 62 |  | 125 |  |
| Observed mean (SD) | 43.5 (23.8) | 185.3 (57.1) |  | 147.2 (56.9) |  | 166.4 (59.9) |  |
| Median (Q1, Q3) | 38 (29, 53) | 177 (155, 210) |  | 137 (114, 160) |  | 157 (128, 197) |  |
| Min, Max | 15, 132 | 93, 405 |  | 64, 343 |  | 64, 405 |  |
| Missing | 0 | 0 |  | 1 |  | 1 |  |
| **Comparison of vaccine preparation time<sup>4</sup>** |  |  |  |  |  |  |  |
| N | 63 | 63 |  | 62 |  | 125 |  |
| LS Mean (95% CI) | 43.5 (31.3, 55.7) | 185.3 (173.1, 197.5) |  | 147.1 (134.9, 159.4) |  | 166.2 (155.9, 176.5) |  |
| LS Mean Difference (95% CI) |  |  | -141.8 (-156.8, -126.7) |  | -103.6 (-118.7, -88.5) |  | -122.7 (-134.2, -111.2) |
| P-value |  |  | p<.0001 |  | p<.0001 |  | p<.0001 |

Abbreviations: CI: confidence interval; LS: least squares Q1, first quartile; Q3, third quartile; SD, standard deviation; VRRs, vaccines that requires reconstitution
<sup>1</sup> Reference
<sup>2</sup> P-value and confidence intervals based on Dunnett multiple comparison adjustment
<sup>3</sup> One participant was unable to participate due to the malfunctioning of a vaccine kit vial adapter
<sup>4</sup> Estimates are derived from the LMM, with sequence and vaccine type as fixed effects, and site and subject as random effects

Adjusting for randomization sequence, vaccine type, site, and individual subject, least square (LS) mean preparation times did not differ from observed means. The LS mean preparation time for PFS was 141.8 seconds (95% CI: 156.8, 126.7; p<0.0001) faster than for VRR1 and 103.6 seconds (118.7, 88.5; p<0.0001) faster than for VRR2 (**Table 2**).

When the two VRRs were pooled together, the mean (95% CI) preparation time was 166.2 seconds (155.9, 176.5). The LS mean preparation time for PFS was 122.7 seconds (95% CI: 134.2, 111.2; p<0.0001) faster than the preparation time for pVRR.

Irrespective of a participant’s prior experience, experience administering vaccines, or occupation, the mean preparation time for PFS was consistently and significantly lower than each individual VRR or pVRR cohort (**Tables S1-S4**).

### Vaccine preparation errors

Vaccine preparation errors were assessed by reviewing the video recordings of each preparation. Agreement on errors between the two observers was 90.5% (n=15). The mean total number of errors (SD) for PFS was 3.5 (1.1), VRR1 was 4.0 (1.5), VRR2 was 3.8 (1.5) and for combined VRRs, 3.9 (1.5). However, the sample size was not large enough to detect differences in errors rates and differences were not statistically significant (**Table 3**). For all three vaccines, QC errors were the most frequently made error type, while only one participant made an asepsis fault error throughout the experiment (for VRR1).

**Table 3.** Vaccine preparation errors, by vaccine and for pooled VRRs.

| Errors | PFS<br>N=63 | VRR1<br>N=63 | VRR2<br>N=63 | pVRRs<br>N=126 |
| --- | --- | --- | --- | --- |
| **Total errors, mean (SD)** | **3.5 (1.1)** | **4.0 (1.5)** | **3.8 (1.5)** | **3.9 (1.5)** |
| **Total quality control errors, mean (SD)** | **1.6 (0.9)** | **2.3 (0.7)** | **1.8 (0.6)** | **2.0 (0.5)** |
| Missed checking expiration date of vaccine kit/vials | 29 (46.0) | 26 (41.3) | 51 (81.0) | 77 (61.1) |
| Missed checking expiration date of syringe | - | 59 (93.7) | - | 59 (46.8) |
| Missed checking expiration date of needle | 57 (90.5) | 59 (93.7) | 60 (95.2) | 119 (94.4) |
| Missed visual inspection | 15 (23.8) | 0 (0) | 2 (3.2) | 2 (1.6) |
| **Total handling errors, mean (SD)** | **0.9 (0.7)** | **1.1 (0.9)** | **1.0 (0.8)** | **1.0 (0.7)** |
| Drop any part of vaccine | 31 (49.2) | 26 (41.3) | 30 (47.6) | 56 (44.4) |
| Lack of gloves | - | 10 (15.9) | - | 10 (7.9) |
| Selected needle of inappropriate size for intramuscular injection | 0 (0) | 13 (20.6) | 11 (17.5) | 24 (19.0) |
| Selected syringe of inappropriate size for aspirating vial contents | 23 (36.5) | 20 (31.7) | 19 (30.2) | 39 (31.0) |
| **Total asepsis fault, mean (SD)** | **0.0 (0.0)** | **0.0 (0.1)** | **0.0 (0.0)** | **0.0 (0.1)** |
| Finger(s) touched vial rubber cap after removal of protective plastic cap | 0 (0) | 0 (0) | 0 (0) | 0 (0) |
| Finger(s) touched top of syringe after removal of protective plastic cap | 0 (0) | 0 (0) | 0 (0) | 0 (0.0) |
| Finger(s) touched sterile part of needle | 0 (0) | 1 (1.6) | 0 (0) | 1 (0.8) |
| Needle stick | 0 (0) | 0 (0) | 0 (0) | 0 (0.0) |
| **Total user technique errors, mean (SD)** | **1.0 (0.4)** | **0.6 (0.7)** | **1.0 (1.1)** | **0.8 (0.8)** |
| Mixing error (vigorous shaking, not swirling, shaking PFS) | 1 (1.6) | 12 (19.0) | 9 (14.3) | 21 (16.7) |
| Dilution occurred | 1 (1.6) | - | - | - |
| Tip cap removal when PFS not upright/removed abruptly/pulled while twisting | 59 (93.7) | - | - | - |
| Purge of syringe at any stage | 0 (0) | - | 0 (0) | 0 (0.0) |
| Lack of cleansing of vial rubber stopper | - | 1 (1.6) | 1 (1.6) | 2 (1.6) |
| Vial content (adjuvant/reconstituted vaccine) not fully aspirated into syringe | - | 1 (1.6) | - | 1 (0.8) |
| Missed reconstitution of vaccine | - | 0 (0) | 1 (1.6) | 1 (0.8) |
| Reconstitution of vaccine using more than one needle (unless damaged/contaminated) | - | 14 (22.2) | 5 (7.9) | 19 (15.1) |
| Spillage or leakage at any stage | 1 (1.6) | 7 (11.1) | 2 (3.2) | 9 (7.1) |
| Attachment of vial adapter at wrong angle | - | - | 20 (31.7) | 20 (15.9) |
| Syringe held at any part but the Luer lock adapter at any stage | - | - | 17 (27.0) | 17 (13.5) |
| Removal of syringe from vial before complete withdrawal after reconstitution | - | - | 4 (6.3) | 4 (3.2) |
| Vial not completely inverted during withdrawal | - | - | 3 (4.8) | 3 (2.4) |
| Plunger rod pulled out of syringe during withdrawal | - | - | 0 (0) | 0 (0.0) |
| **Other** | **1 (1.6)** | **3 (4.8)** | **4 (6.3)** | **7 (5.6)** |
| **Missing** | **1 (1.6)** | **3 (4.8)** | **4 (6.3)** | **7 (5.6)** |

Note: Based on Poisson GLMM comparing preparation errors, with sequence and vaccine type as fixed effects, and site and subject as random effects, incidence rate ratio (95% CI; p-value): PFS vs VRR1 [0.87 (0.72, 1.04; p=0.1197)]; PFS vs VRR2 [0.92 (0.76, 1.10; p=0.3559)]; PFS vs pVRRs [0.89 (0.76, 1.05; p= 0.1571)]

### Vaccine satisfaction and preference

Participants frequently indicated that they were ‘Very Satisfied’ or ‘Extremely Satisfied’ with the PFS vaccine in terms of the ease-of preparation (n=12 and 43, respectively; 87.3% total) and preparation time (n=7 and 49, respectively; 88.9% total) (**Figure 3** and **Table S5**). Satisfaction scores for VRR1 and VRR2 were more widely distributed, and the most selected satisfaction score for both vaccines was ‘Satisfied’ for each satisfaction measure. Overall, only 1.6% (n=1) of participants indicated any degree of dissatisfaction (combined ‘Dissatisfied,’ ‘Very Dissatisfied,’ and ‘Extremely Dissatisfied’) for PFS, while 20.6% and 17.5% said the same for VRR1 (n=13) and VRR2 (n=11), respectively. The overall satisfaction (combined ‘Very Satisfied’ and ‘Extremely Satisfied’) was 87.3% (n=55) for PFS, 28.6% (n=18) for VRR1, and 47.6% (n=30) for VRR2 (**Figure 3** and **Table S5**).

**Figure 3.**
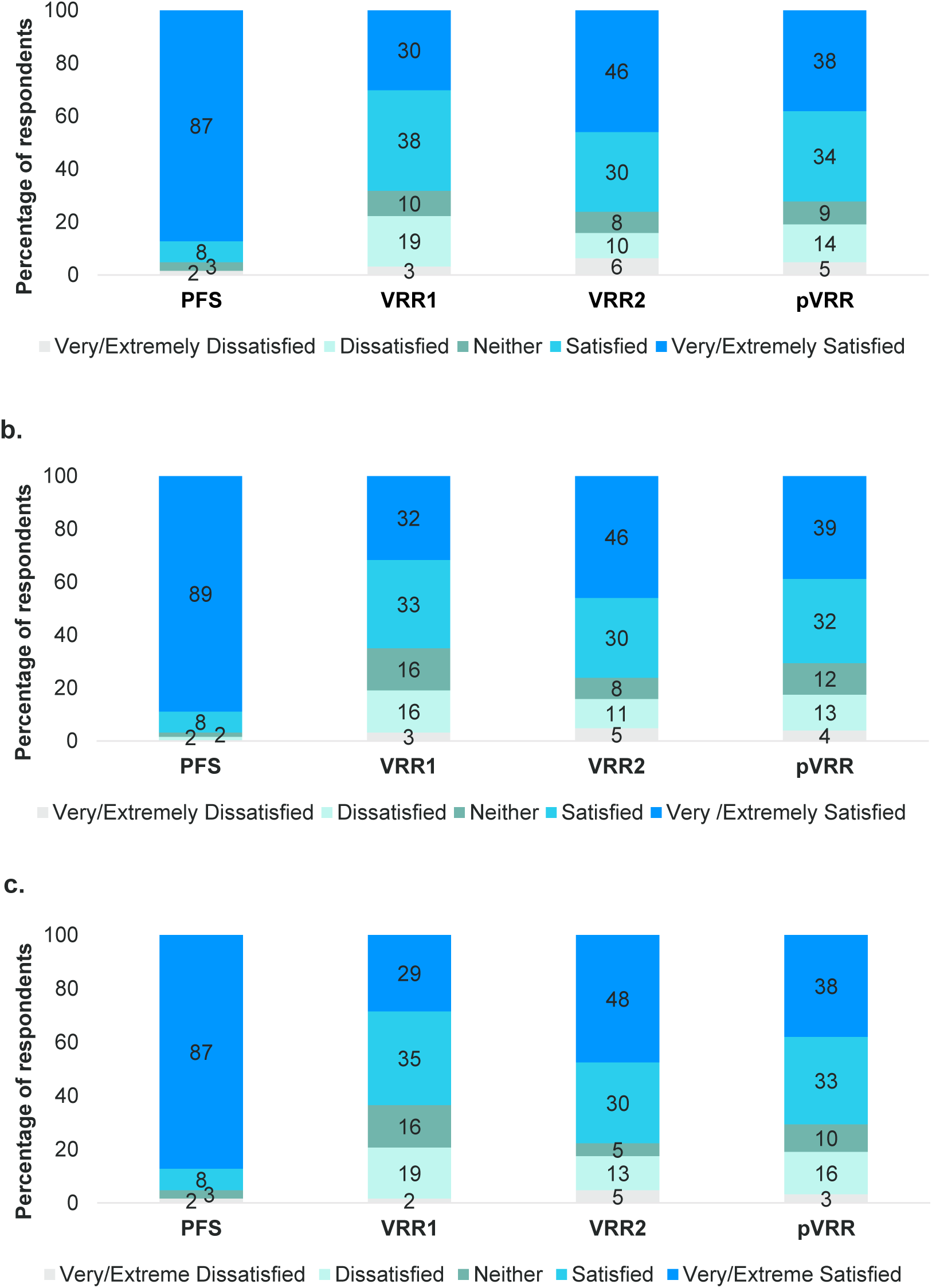
Participant Satisfaction Ratings for a) Ease of preparation, b) Time of preparation, c) Overall satisfaction with vaccine preparation.

Compared to VRR1 and VRR2, PFS had 19.93 (95% CI: 8.32, 47.72; p<0.0001) and 10.87 (95% CI: 4.72, 25.06; p<0.0001) times higher odds, respectively, of receiving a higher overall satisfaction score. Similarly, compared to VRRs combined, PFS had 14.72 (95% CI: 6.68, 32.41; p<0.0001) times greater odds of receiving a higher overall satisfaction score. This trend was consistent across ‘ease of use’ and ‘preparation time’ satisfaction measures (**Table 4**).

**Table 4.** Comparison of participant satisfaction with vaccine preparation procedures.

|  | Odds Ratios (95% CI) | p-value |
| --- | --- | --- |
| **Satisfaction with Ease of Use** |  |  |
| PFS vs. VRR1 | 18.41 (7.70, 43.99) | <.0001 |
| PFS vs. VRR2 | 10.61 (4.61, 24.37) | <.0001 |
| PFS vs. pVRRs | 13.97 (6.37, 30.66) | <.0001 |
| **Satisfaction with Preparation Time** |  |  |
| PFS vs. VRR1 | 34.60 (12.85, 93.16) | <.0001 |
| PFS vs. VRR2 | 20.16 (7.78, 52.20) | <.0001 |
| PFS vs. pVRRs | 26.41 (10.60, 65.82) | <.0001 |
| **Overall Satisfaction** |  |  |
| PFS vs. VRR1 | 19.93 (8.32, 47.72) | <.0001 |
| PFS vs. VRR2 | 10.87 (4.72, 25.06) | <.0001 |
| PFS vs. pVRRs | 14.72 (6.68, 32.41) | <.0001 |

Abbreviations: CI: confidence interval Estimates derived from a mixed-effects ordered logit regression models, with sequence and vaccine type as fixed effects, and site and subject as random effects

After all vaccine preparations were complete, participants selected their preferred vaccine. The majority of participants (81.0%) reported a preference for the ready-to-use prefilled syringe vaccine, PFS (**Figure 4** and **Table S6**).

**Figure 4.**
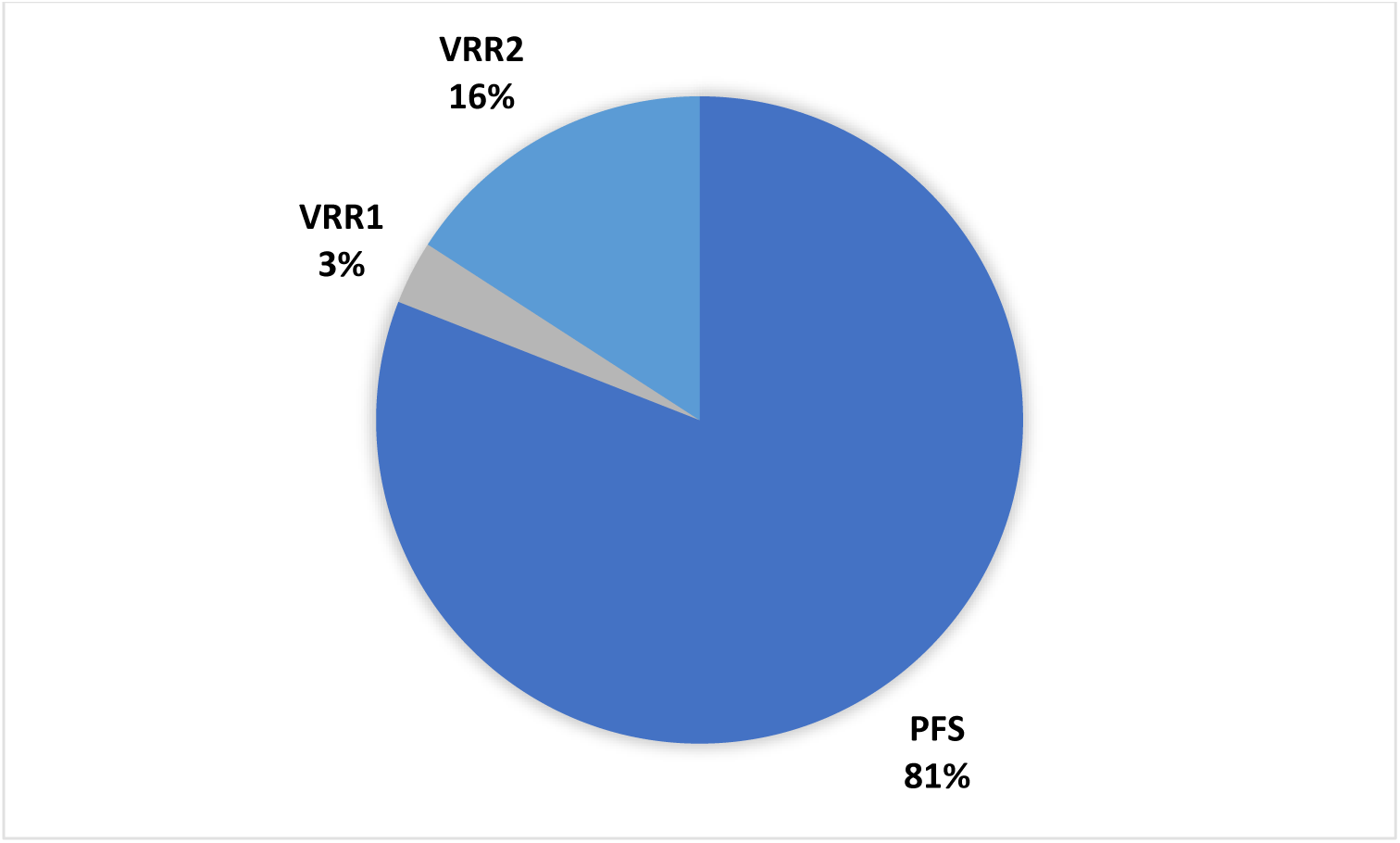
Participant preference for vaccine preparations.

This is the first study to compare vaccine preparation time, errors, satisfaction, and preferences for RSV vaccines. We found that preparation time with PFS was significantly lower as compared to VRRs, both when testing each VRR separately and when combining them. Study participants were extremely satisfied and preferred a PFS over VRRs. We did not observe any significant differences in the number of errors between vaccine preparations.

### Vaccine preparation time

Consistent with prior published literature [11, 13], results from the study demonstrated that the preparation time with a PFS vaccine was nearly 4 times lower as compared to vaccines that require reconstitution. The preparation time with PFS was significantly lower as compared to VRR1 and VRR2, both when testing each VRR separately and when combining them. Based on the observed vaccine preparation times, we can infer that vaccine administrators can prepare 83 PFS vaccines per hour, 20 VRR1 vaccines per hour, and 24 VRR2 vaccines per hour consecutively. Considering the timing of pVRR, we can infer that vaccine administrators can prepare 22 reconstituted vaccines per hour.

Of note, recent updates have been made to the label for VRR2 (ABRYSVO™), which now includes two presentations; a vial of lyophilized powder and a vial of liquid diluent, and a vial of lyophilized powder and a PFS diluent [9], the latter of which was used in this study. Although both require reconstitution, use of the vial and PFS presentation may explain the slightly lower preparation times compared to VRR1. The vaccine kit for VRR1 includes two vials, and preparation steps are therefore more like those of the VRR2 vial and vial presentation.

The majority of the population receives their vaccines in a pharmacy setting [21, 22]. Due to the start of respiratory season and recommendations guiding the administration of RSV and influenza vaccines, are administered during the fall season every year [7] resulting in increased pressure on community pharmacies resources. A recent survey of community pharmacists in the US has demonstrated that 74.9% of the pharmacists were experiencing burnout [23–26]. Further, a systematic review identified 19 articles across eight countries and estimated a burnout prevalence of 51% [23]. Some of the risk factors associated with burnout were high patient and prescription volumes and increased workload. The increased demand for vaccination during the fall season might result in higher patient volume and workload leading to pharmacists’ burnout and potential errors [27, 28]. Results from the study suggest that an alternative RSV vaccine in a ready-to-use PFS might reduce the heavy workload that leads to pharmacist burnout.

### Vaccine preparation errors

The total number of errors made during preparation did not differ significantly between vaccines. Likewise, the total number of vaccine-specific errors were similar across vaccines. The ability to detect statistical differences in error rates in our study was limited because the study was powered for the primary outcome detecting time differences and not for error rates. Further, this was a controlled experiment and error rates in a busy retail clinic may not be accurately reflected here.

For the PFS vaccine, user technique error related to tip cap removal was the main source of preparation errors. These errors did not result in any leaking of the vaccine and hence would not impact product quality or prevent vaccine administration. For the VRR1 and VRR2, there was a numerical trend for higher proportion of spillage or leakage. In addition, due to the requirement for reconstitution and additional associated steps, additional errors such as ‘Lack of cleansing of vials rubber stopper after removing the plastic flip off’ or ‘Vial content (adjuvant or reconstituted vaccine) not fully aspired into syringe’ were observed. This is in line with other published studies [13, 16]. A survey-based evaluation of vaccine related errors by Lee et al. found 76.4% of physicians and 41.5% of nurses experienced errors related to reconstitution, including ‘vial contents not aspired into syringe’ (52.0% and 14.7%, respectively), ‘inadequate shaking’ (51.6% and 19.1%, respectively), ‘spillage or leakage during reconstitution’ (42.4% and 14.7%, respectively), ‘needle twisted when inserted in vial stopper’ (29.6% and 11.8%, respectively), ‘same needle used for reconstitution and injection’ (23.2% and 10.0%, respectively), ‘forgetting to reconstitute’ (19.2% and 7.6%, respectively), ‘other’ (31.6% and 11.1%, respectively) [16].

### Satisfaction and preferences

In line with previous published literature, the study results showed that study participants were satisfied and preferred a PFS over VRRs. Though understanding the drivers of this satisfaction and preference were out of the study scope, the high satisfaction on ease and speed of preparation indicate these are contributing factors to the overall preference. In support, a recent targeted literature review found PFS was most often preferred by physicians and nurses with common reasons cited that included reduced immunization errors, errors typically occurring during reconstitution, followed by ease of administration [12].

### Study limitations

There are several limitations to the current study design, including the inability to completely blind observers to the three vaccines given the differences between preparations, and it is possible that the observers were familiar with the different vaccines. While the strength of the cross-over design allowed for subjects to act as their own controls, the order in which the vaccines were prepared may have affect outcomes including timing, errors, satisfaction, and preference. To minimize the effects of order, we utilized a 3-sequence block design to balance the order of vaccine preparation among participants. Lastly, participants performed a single assessment for each vaccine whereas if repeated preparations were performed there may have been trained efficiencies gained.

The strengths of the design lie in the direct time assessment and recorded review of preparations that allowed for repeated views to determine timing and errors. In addition, independent assessments between observers were performed to ensure agreement (>80%).

## Conclusions

Given the workload in pharmacy settings, implementation of an RSV program using a PFS vaccine may provide significant advantages. PFS vaccines can greatly simplify the vaccine preparation process, allowing administrators to prepare almost four times more doses per hour than with vial and syringe systems.

## Acknowledgements

We would like to thank the following people: Anna Krivelyova (ICON Plc) for overall project support; Mohana Giruparajah and Adeola Adejumo (ICON Plc) for their operational support to the project; Jaya Paranilam (ICON Plc) for the IRB submission; Ruchika Sharma and Shravan Kumar Adepu (ICON Plc) for running the statistical analysis; Natalia Olter and Sabine Grothues (ICON Plc) for reviewing the vaccine preparation video-recordings for time and errors; Kathleen Walker for coordinating the AccellaCare sites; Katy Klein, Erin Hackett, Emma Hughes, Devon Brown, Djemila Fields, Chernessa Campbell, Sarah Utech, and Alphonso Chambers (AccellaCare) for coordinating participants and data collection at site level; Erin Butler, Jennifer Haynes, Sydney Rabinowitz, and Tammy Joseph (GLG) for recruitment of participants; all healthcare professional who participated to the study preparing the vaccines and completing the survey for their time.

## Author Contributions

DM, SK, and MM were involved in study conception. Material preparation was performed by SF, DC, MM, and SK. All authors were involved in the study design. Data analysis was performed by MH and AP. The manuscript was drafted by SF, DC, and SK. All authors commented on drafts of the manuscript and read and approved the final manuscript.

## Ethics/Ethical Approval

This study received Institutional Review Board (IRB) approval from Advarra Central IRB (MOD01999449).

## Data Availability

All data generated during and/or analyzed during the current study are available from the corresponding author on reasonable request.

## Supplementary Material 1: Preference and Satisfaction Survey

1. What is your gender? (Male/Female/Non-binary/Other)
2. What is your age range? years
3. What is your occupation? (Pharmacist/Nurse/Pharmacy technician/Other *(open field)*)
4. How would you describe the area in which you practice? Check all that apply. (Urban/Rural/Suburban)
5. How would you describe the setting in which you practice? Check all that apply. (Hospital/Clinical pharmacy/Academic institution/Retail pharmacy/Other *(open field)*)
6. How many years’ experiences you have as a practicing pharmacist/certified nurse/pharmacy technician? (1-10 years; 11-20 years; 21-30 years; 31-40 years; >40 years)
7. How many years have you been administering vaccines? (1.5-3 years; >3-5 years; >5-10 years; >10 years)
8. What is the average number of vaccines you administer in a typical month? (1-10/11- 20/21-30/31-40/40+ vaccines/month)
9. Have you received training on vaccine preparation in general? (Yes/No)
10. Have you received training on vaccine preparation specific for vaccine in a ready-to-use prefilled syringe? (Yes/No)
11. Have you ever prepared a vaccine in a ready-to-use prefilled syringe before? (Yes/No)
12. Have you received training on vaccine preparation specific for vaccines that require reconstitution? (Yes/No)
13. Have you ever prepared a vaccine that requires reconstitution before? (Yes/No)

### Stop! Please answer the next section after completing the preparation of the first vaccine

1. Which vaccine did you just prepare? (A/B/C)
2. How satisfied or dissatisfied are you with the vaccine ease-of-preparation?

□ Extremely Dissatisfied
□ Very Dissatisfied
□ Dissatisfied
□ Neither satisfied nor dissatisfied
□ Satisfied
□ Very Satisfied
□ Extremely Satisfied
3. How satisfied or dissatisfied are you with the time it took to prepare the vaccine?

□ Extremely Dissatisfied
□ Very Dissatisfied
□ Dissatisfied
□ Neither satisfied nor dissatisfied
□ Satisfied
□ Very Satisfied
□ Extremely Satisfied
4. Taking all things into account, how satisfied or dissatisfied are you overall with the vaccine preparation procedure?

□ Extremely Dissatisfied
□ Very Dissatisfied
□ Dissatisfied
□ Neither satisfied nor dissatisfied
□ Satisfied
□ Very Satisfied
□ Extremely Satisfied
5. Would you feel comfortable administering this preparation to a patient?

□ Yes
□ No
If the answer to 18 is No, the follow question appears:
Please indicate the reason:
□ Particulate matter inside the syringe
□ Appearance issue (of the content itself or of the syringe or tip cap being discolored)
□ Broken/cracked syringe
□ Label issue (missing information, illegible, missing label upon opening of the salespack)
□ Hard to remove the tip cap
□ Plunger rod issues (falling off, not present)
□ Issues with the volume inside the syringe (too high, too low, empty)
□ Leaking
□ Other, please specify: [free text]

### Stop! Please answer the next section after completing the preparation of the second vaccine

1. Which vaccine did you just prepare? (A/B/C)
2. How satisfied or dissatisfied are you with the vaccine ease-of-preparation?

□ Extremely Dissatisfied
□ Very Dissatisfied
□ Dissatisfied
□ Neither satisfied nor dissatisfied
□ Satisfied
□ Very Satisfied
□ Extremely Satisfied
3. How satisfied or dissatisfied are you with the time it took to prepare the vaccine?

□ Extremely Dissatisfied
□ Very Dissatisfied
□ Dissatisfied
□ Neither satisfied nor dissatisfied
□ Satisfied
□ Very Satisfied
□ Extremely Satisfied
4. Taking all things into account, how satisfied or dissatisfied are you overall with the vaccine preparation procedure?

□ Extremely Dissatisfied
□ Very Dissatisfied
□ Dissatisfied
□ Neither satisfied nor dissatisfied
□ Satisfied
□ Very Satisfied
□ Extremely Satisfied
5. Would you feel comfortable administering this preparation to a patient?

□ Yes
□ No
If the answer to 23 is No, the follow question appears:
Please indicate the reason:
□ Particulate matter inside the syringe
□ Appearance issue (of the content itself or of the syringe or tip cap being discolored)
□ Broken/cracked syringe
□ Label issue (missing information, illegible, missing label upon opening of the salespack)
□ Hard to remove the tip cap
□ Plunger rod issues (falling off, not present)
□ Issues with the volume inside the syringe (too high, too low, empty)
□ Leaking
□ Other, please specify: [free text]

#### Stop! Please answer the remainder of the survey after completing the preparation of the final vaccine

1. Which vaccine did you just prepare? (A/B/C)
2. How satisfied or dissatisfied are you with the vaccine ease-of-preparation?

□ Extremely Dissatisfied
□ Very Dissatisfied
□ Dissatisfied
□ Neither satisfied nor dissatisfied
□ Satisfied
□ Very Satisfied
□ Extremely Satisfied
3. How satisfied or dissatisfied are you with the time it took to prepare the vaccine?

□ Extremely Dissatisfied
□ Very Dissatisfied
□ Dissatisfied
□ Neither satisfied nor dissatisfied
□ Satisfied
□ Very Satisfied
□ Extremely Satisfied
4. Taking all things into account, how satisfied or dissatisfied are you overall with the vaccine preparation procedure?

□ Extremely Dissatisfied
□ Very Dissatisfied
□ Dissatisfied
□ Neither satisfied nor dissatisfied
□ Satisfied
□ Very Satisfied
□ Extremely Satisfied
5. Would you feel comfortable administering this preparation to a patient?

□ Yes
□ No
If the answer to 28 is No, the follow question appears:
Please indicate the reason:
□ Particulate matter inside the syringe
□ Appearance issue (of the content itself or of the syringe or tip cap being discolored)
□ Broken/cracked syringe
□ Label issue (missing information, illegible, missing label upon opening of the salespack)
□ Hard to remove the tip cap
□ Plunger rod issues (falling off, not present)
□ Issues with the volume inside the syringe (too high, too low, empty)
□ Leaking
□ Other, please specify: [free text]
6. Which one of the three vaccine preparation processes do you prefer the most?

*Tick the option stating the letter of the blinded bottle*
*Question 26 is the last question, and the order of the possible answers in it will be randomized.*
□ Vaccine A
□ Vaccine B
□ Vaccine C

## Supplementary Material 2: Errors List

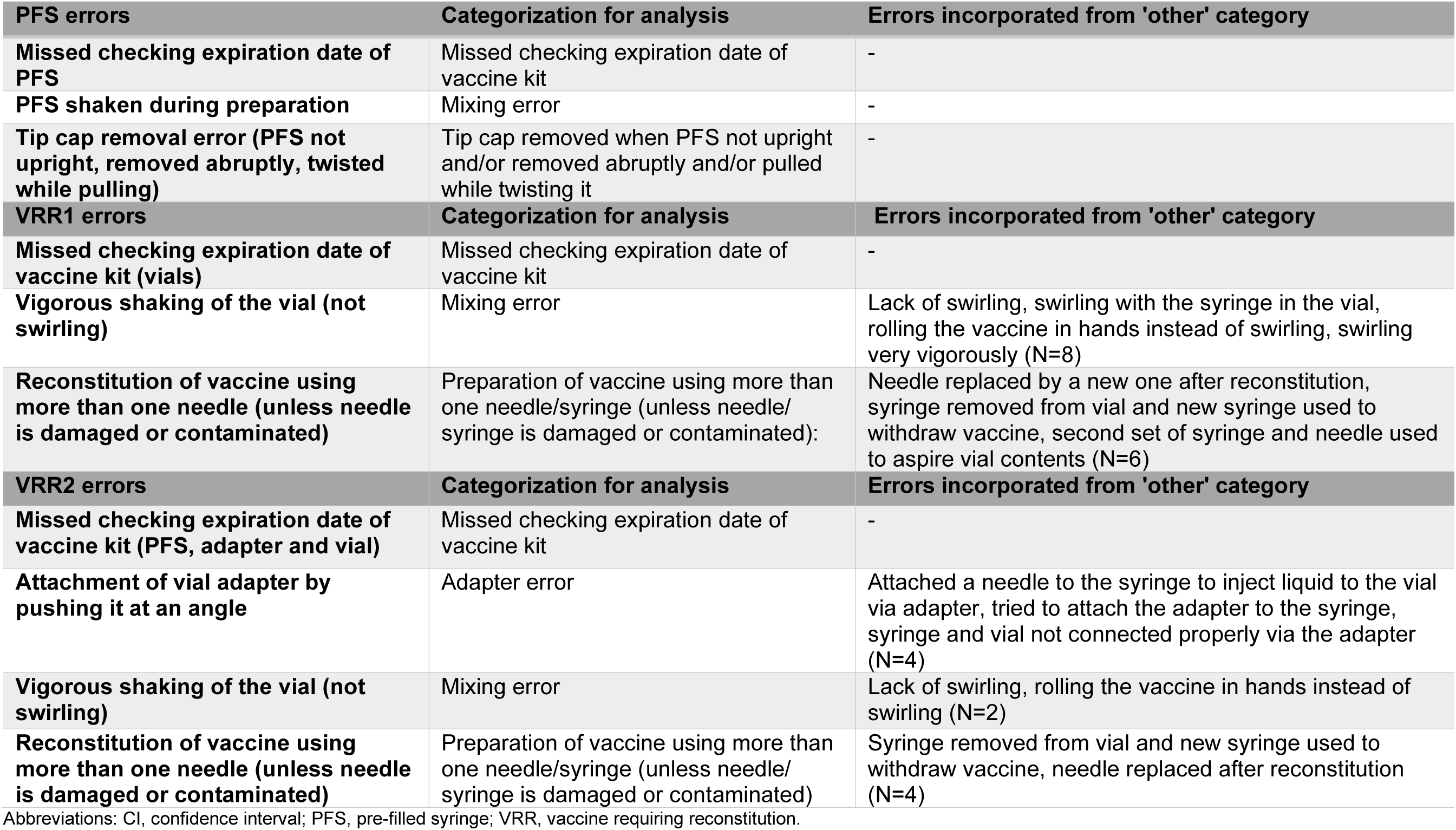

| PFS errors | Categorization for analysis | Errors incorporated from 'other' category |
| --- | --- | --- |
| Missed checking expiration date of PFS | Missed checking expiration date of vaccine kit | - |
| PFS shaken during preparation | Mixing error | - |
| Tip cap removal error (PFS not upright, removed abruptly, twisted while pulling) | Tip cap removed when PFS not upright and/or removed abruptly and/or pulled while twisting it | - |
| VRR1 errors | Categorization for analysis | Errors incorporated from 'other' category |
| Missed checking expiration date of vaccine kit (vials) | Missed checking expiration date of vaccine kit | - |
| Vigorous shaking of the vial (not swirling) | Mixing error | Lack of swirling, swirling with the syringe in the vial, rolling the vaccine in hands instead of swirling, swirling very vigorously (N=8) |
| Reconstitution of vaccine using more than one needle (unless needle is damaged or contaminated) | Preparation of vaccine using more than one needle/syringe (unless needle/syringe is damaged or contaminated): | Needle replaced by a new one after reconstitution, syringe removed from vial and new syringe used to withdraw vaccine, second set of syringe and needle used to aspire vial contents (N=6) |
| VRR2 errors | Categorization for analysis | Errors incorporated from 'other' category |
| Missed checking expiration date of vaccine kit (PFS, adapter and vial) | Missed checking expiration date of vaccine kit | - |
| Attachment of vial adapter by pushing it at an angle | Adapter error | Attached a needle to the syringe to inject liquid to the vial via adapter, tried to attach the adapter to the syringe, syringe and vial not connected properly via the adapter (N=4) |
| Vigorous shaking of the vial (not swirling) | Mixing error | Lack of swirling, rolling the vaccine in hands instead of swirling (N=2) |
| Reconstitution of vaccine using more than one needle (unless needle is damaged or contaminated) | Preparation of vaccine using more than one needle/syringe (unless needle/syringe is damaged or contaminated) | Syringe removed from vial and new syringe used to withdraw vaccine, needle replaced after reconstitution (N=4) |

Abbreviations: CI, confidence interval; PFS, pre-filled syringe; VRR, vaccine requiring reconstitution.

## Supplementary Material 3: Errors included in ‘other’

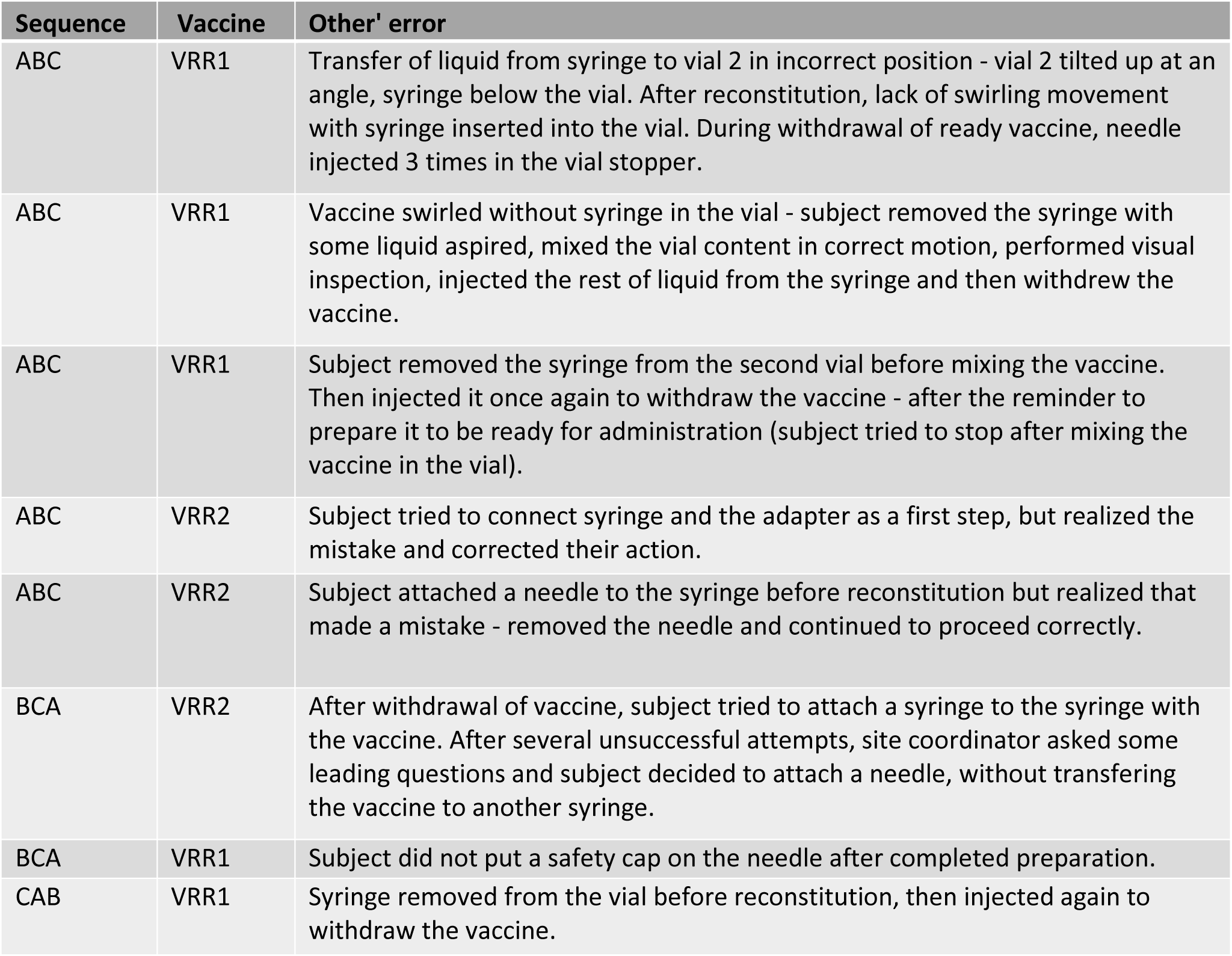
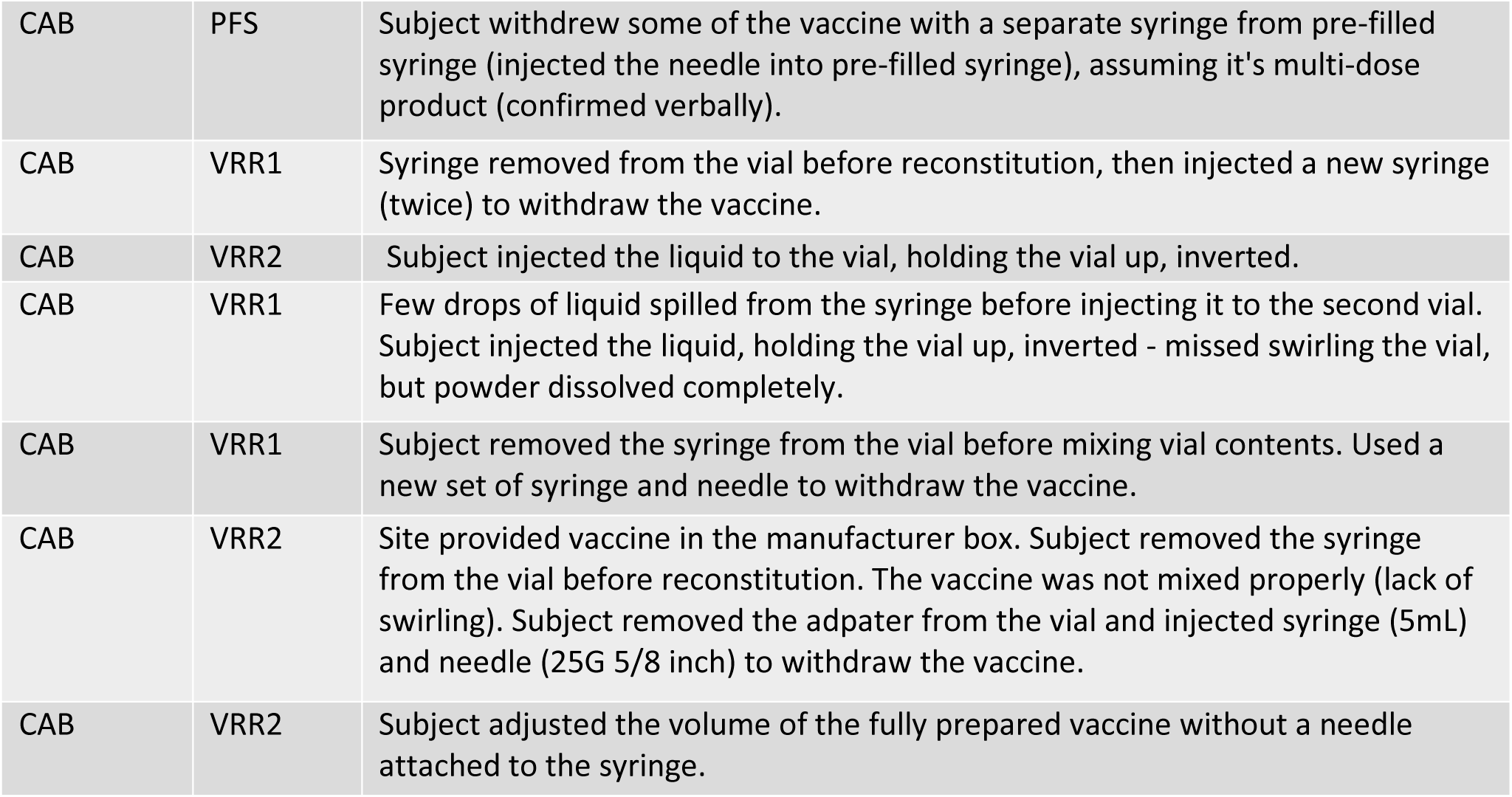

| Sequence | Vaccine | Other' error |
| --- | --- | --- |
| ABC | VRR1 | Transfer of liquid from syringe to vial 2 in incorrect position - vial 2 tilted up at an angle, syringe below the vial. After reconstitution, lack of swirling movement with syringe inserted into the vial. During withdrawal of ready vaccine, needle injected 3 times in the vial stopper. |
| ABC | VRR1 | Vaccine swirled without syringe in the vial - subject removed the syringe with some liquid aspired, mixed the vial content in correct motion, performed visual inspection, injected the rest of liquid from the syringe and then withdrew the vaccine. |
| ABC | VRR1 | Subject removed the syringe from the second vial before mixing the vaccine. Then injected it once again to withdraw the vaccine - after the reminder to prepare it to be ready for administration (subject tried to stop after mixing the vaccine in the vial). |
| ABC | VRR2 | Subject tried to connect syringe and the adapter as a first step, but realized the mistake and corrected their action. |
| ABC | VRR2 | Subject attached a needle to the syringe before reconstitution but realized that made a mistake - removed the needle and continued to proceed correctly. |
| BCA | VRR2 | After withdrawal of vaccine, subject tried to attach a syringe to the syringe with the vaccine. After several unsuccessful attempts, site coordinator asked some leading questions and subject decided to attach a needle, without transferring the vaccine to another syringe. |
| BCA | VRR1 | Subject did not put a safety cap on the needle after completed preparation. |
| CAB | VRR1 | Syringe removed from the vial before reconstitution, then injected again to withdraw the vaccine. |
| CAB | PFS | Subject withdrew some of the vaccine with a separate syringe from pre-filled syringe (injected the needle into pre-filled syringe), assuming it's multi-dose product (confirmed verbally). |
| CAB | VRR1 | Syringe removed from the vial before reconstitution, then injected a new syringe (twice) to withdraw the vaccine. |
| CAB | VRR2 | Subject injected the liquid to the vial, holding the vial up, inverted. |
| CAB | VRR1 | Few drops of liquid spilled from the syringe before injecting it to the second vial. Subject injected the liquid, holding the vial up, inverted - missed swirling the vial, but powder dissolved completely. |
| CAB | VRR1 | Subject removed the syringe from the vial before mixing vial contents. Used a new set of syringe and needle to withdraw the vaccine. |
| CAB | VRR2 | Site provided vaccine in the manufacturer box. Subject removed the syringe from the vial before reconstitution. The vaccine was not mixed properly (lack of swirling). Subject removed the adapter from the vial and injected syringe (5mL) and needle (25G 5/8 inch) to withdraw the vaccine. |
| CAB | VRR2 | Subject adjusted the volume of the fully prepared vaccine without a needle attached to the syringe. |

## Supplementary Tables

**Table S1.** Comparison of vaccine preparation time - supplementary model 1 (adjusting for experience in occupation)

| Least-Square Mean (95% CI) |  |  | Type III test results |  | Least-Square Mean Differences (95% CI), p value |  |  |  |
| --- | --- | --- | --- | --- | --- | --- | --- | --- |
| PFS | VRR1 | VRR2 | F-Value | p-value | PFS vs. VRR1 |  | PFS vs. VRR2 |  |
| 43.6 (31.5, 55.6) | 185.3 (173.3, 197.4) | 147.1 (135.0, 159.3) | 238.75 | <.0001 | -141.8 (-155.0, -128.5) | p=<.0001 | -103.6 (-116.9, -90.2) | p=<.0001 |
| Regression coefficients for experience in occupation (Ref: 1-10 years) |  |  | F-Value | p-value |  |  |  |  |
| 1-10 years | - | - | 1.58 | 0.1843 | - |  | - |  |
| 11-20 year | -10.9 (-33.1, 11.2) | p=0.3292 |  |  | - |  | - |  |
| 21-30 year | 13.9 (-15.2, 42.9) | p=0.3457 |  |  | - |  | - |  |
| 31-40 year | 1.7 (-38.4, 41.9) | p=0.9321 |  |  | - |  | - |  |
| >40 years | 33.9 (-6.1, 73.8) | p=0.0958 |  |  | - |  | - |  |

Abbreviations: CI, confidence interval; PFS, pre-filled syringe; VRR, vaccine requiring reconstitution.
Estimates are derived from the LMM, with sequence, vaccine and experience in occupation as fixed effects, and site and subject as random effects

**Table S2.** Comparison of vaccine preparation time - supplementary model 2 (adjusting for experience administering vaccines)

| Least-Square Mean (95% CI) |  |  | Type III test results |  | Least-Square Mean Differences (95% CI), p value |  |  |  |
| --- | --- | --- | --- | --- | --- | --- | --- | --- |
| PFS | VRR1 | VRR2 | F-Value | p-value | PFS vs. VRR1 |  | PFS vs. VRR2 |  |
| 43.5 (31.6, 55.4) | 185.3 (173.4, 197.1) | 147.1 (135.2, 159.1) | 238.83 | <.0001 | -141.8 (-155.0, -128.5) | p=<.0001 | -103.6 (-117.0, -90.3) | p=<.0001 |
| Regression coefficients for experience administering vaccines (Ref: 1.5-3 years) |  |  | F-Value | p-value |  |  |  |  |
| 1.5-3 years |  |  | 2.97 | 0.0345 | - |  | - |  |
| >3-5 years | -50.6 (-84.9, -16.4) | p=0.0040 |  |  | - |  | - |  |
| >5-10 years | -20.5 (-52.9, 11.9) | p=0.2126 |  |  | - |  | - |  |
| >10 years | -24.8 (-53.7, 4.0) | p=0.0907 |  |  | - |  | - |  |

Abbreviations: CI, confidence interval; PFS, pre-filled syringe; VRR, vaccine requiring reconstitution.
Estimates are derived from the LMM, with sequence, vaccine and experience administering vaccines as fixed effects, and site and subject as random effects

**Table S3.** Comparison of vaccine preparation time - supplementary model 3 (adjusting for occupation)

| Least-Square Mean (95% CI) |  |  | Type III test results |  | Least-Square Mean Differences (95% CI), p value |  |  |  |
| --- | --- | --- | --- | --- | --- | --- | --- | --- |
| PFS | VRR1 | VRR2 | F-Value | p-value | PFS vs. VRR1 |  | PFS vs. VRR2 |  |
| 43.5 (31.5, 55.6) | 185.3 (173.2, 197.4) | 147.1 (135.0, 159.3) | 238.78 | <.0001 | -141.8 (-155.0, -128.5) | p=<.0001 | -103.6 (-116.9, -90.2) | p=<.0001 |
| Regression coefficients for occupation (Ref: Pharmacist) |  |  | F-Value | p-value |  |  |  |  |
| Pharmacist | - | - | 2.13 | 0.1237 | - |  | - |  |
| Pharmacy technician | -10.3 (-55.5, 34.8) | p=0.6511 |  |  | - |  | - |  |
| Nurse | 19.2 (-0.7, 39.1) | p=0.0588 |  |  | - |  | - |  |

Abbreviations: CI, confidence interval; PFS, pre-filled syringe; VRR, vaccine requiring reconstitution.
Estimates are derived from the LMM, with sequence, vaccine and occupation as fixed effects, and site and subject as random effects

**Table S4.** Comparison of vaccine preparation time - post-hoc analyses of mRNA-1273 vs. VRRs.

| Model | Least-Square Mean (95% CI) |  | Least-Square Mean Differences (95% CI),<br>p value |  |
| --- | --- | --- | --- | --- |
|  | PFS | pVRR | PFS vs. pVRR |  |
| Supplementary model 1: Adjusting for experience in occupation | 43.6 (31.5, 55.6) | 166.2 (156.1, 176.3) | -122.7 (-134.2, -111.1) | p<.0001 |
| Supplementary model 2: Adjusting for experience administering vaccines | 43.5 (31.6, 55.4) | 166.2 (156.3, 176.1) | -122.7 (-134.2, -111.2) | p<.0001 |
| Supplementary model 3: Adjusting for occupation | 43.5 (31.5, 55.6) | 166.2 (156.1, 176.3) | -122.7 (-134.2, -111.2) | p<.0001 |

Abbreviations: CI, confidence interval; PFS, pre-filled syringe; pVRR, pulled vaccines requiring reconstitution.
Estimates are derived from the LMM, with sequence, vaccine, and the respective covariate as fixed effects, and site and subject as random effects

**Table S5.**
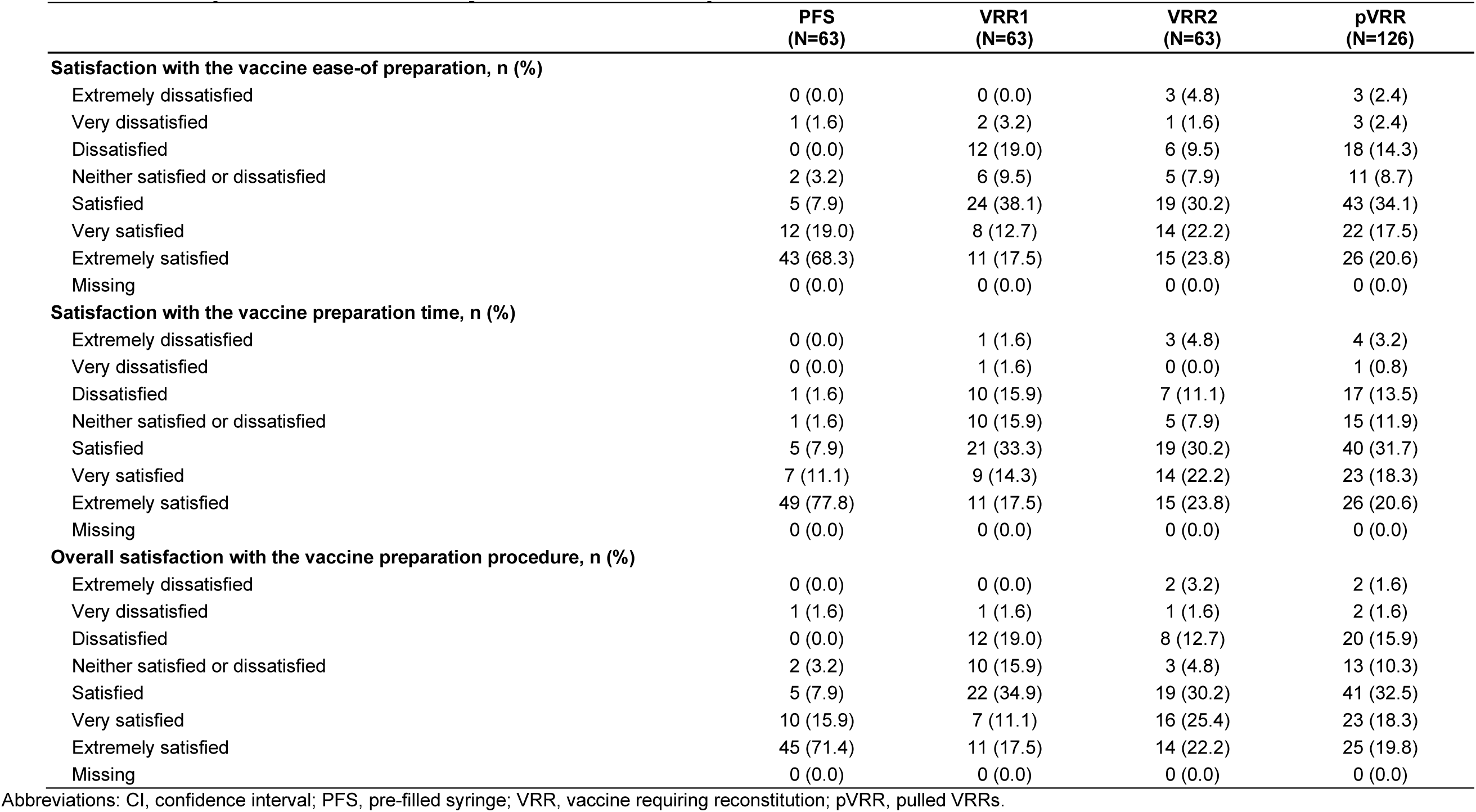
Participant satisfaction, by vaccine and for pooled VRRs.

**Table S6.** Participant preference of vaccine.

|  | (N=63) |
| --- | --- |
| **Preferred Vaccine** |  |
| PFS | 51 (81.0) |
| VRR1 | 2 (3.2) |
| VRR2 | 10 (15.9) |
| Missing | 0 (0.0) |

Abbreviations: CI, confidence interval; PFS, pre-filled syringe; VRR, vaccine requiring reconstitution.

